# OpenABM-Covid19 - an agent-based model for non-pharmaceutical interventions against COVID-19 including contact tracing

**DOI:** 10.1101/2020.09.16.20195925

**Authors:** Robert Hinch, William J M Probert, Anel Nurtay, Michelle Kendall, Chris Wymant, Matthew Hall, Katrina Lythgoe, Ana Bulas Cruz, Lele Zhao, Andrea Stewart, Luca Ferretti, Daniel Montero, James Warren, Nicole Mather, Matthew Abueg, Neo Wu, Anthony Finkelstein, David G Bonsall, Lucie Abeler-Dörner, Christophe Fraser

**Affiliations:** Big Data Institute, Li Ka Shing Centre for Health Information and Discovery, Nuffield Department of Medicine, University of Oxford, Oxford, UK; IBM United Kingdom, Portsmouth, UK; Google Research, Mountain View, CA, USA; Department of Computer Science, University College London, London, UK, and Alan Turing Institute, London, UK; Oxford University NHS Trust, University of Oxford, Oxford, UK; Wellcome Centre for Human Genetics, University of Oxford, Oxford, UK

**Author notes:** these authors contributed equally to this work (Robert Hinch, William J M Probert).

## Abstract

SARS-CoV-2 has spread across the world, causing high mortality and unprecedented restrictions on social and economic activity. Policymakers are assessing how best to navigate through the ongoing epidemic, with models being used to predict the spread of infection and assess the impact of public health measures. Here, we present OpenABM-Covid19: an agent-based simulation of the epidemic including detailed age-stratification and realistic social networks. By default the model is parameterised to UK demographics and calibrated to the UK epidemic, however, it can easily be re-parameterised for other countries. OpenABM-Covid19 can evaluate non-pharmaceutical interventions, including both manual and digital contact tracing. It can simulate a population of 1 million people in seconds per day allowing parameter sweeps and formal statistical model-based inference. The code is open-source and has been developed by teams both inside and outside academia, with an emphasis on formal testing, documentation, modularity and transparency. A key feature of OpenABM-Covid19 is its Python interface, which has allowed scientists and policymakers to simulate dynamic packages of interventions and help compare options to suppress the COVID-19 epidemic.

## 1. Introduction

The novel coronavirus SARS-CoV-2 first appeared in China in late 2019 and spread across the globe in early 2020, causing several hundred thousand deaths world-wide in the first half of the year and overwhelming health systems [1]. Restrictions on movement were imposed in many countries, with severe impacts on social life, education, and economies [2]. Mathematical models have long been used to explain and forecast the course of epidemics and to predict the effects of public health interventions [3,4]. Most governments and policymakers use mathematical models to inform their decision-making [5]. The scientific community has responded by adapting old models and designing new models to learn more about the COVID-19 epidemic and inform public health.

Compared to compartmental models and branching-process models, agent-based models of the spread of infection allow for a more complete representation of the social contact network in which contagion occurs [6]. Major advantages include the ability to model heterogeneity in contact rates and local saturation effects, and the ability to better model contact tracing. Alongside other non-pharmaceutical interventions, contact tracing is an important intervention to help reduce the spread of COVID-19 [7,8]. In an agent-based model, the full history of all contacts can be stored, allowing for the impact of contact tracing to be explored in detail. For example, agent-based models can include clustering in the contact network, so if incidence is high in a region of the contact network, an uninfected person who is contract-traced will be protected from this high level of local incidence. A downside of agent-based models is that they are comparatively complex to code, are often not very parsimonious, and can be very computationally intensive to run, limiting the ability to explore a wide range of parameter combinations. Here, we focus on developing OpenABM-Covid19, an agent-based simulation which addresses these downsides, by focussing on parsimony, computational efficiency, code transparency, and a robust testing framework.

A particular focus of our work applying OpenABM-Covid19 has been exploring different ways in which contact tracing, and in particular digital contact tracing using mobile phone apps that record proximity events, can contribute to epidemic control [9]. Several other groups have approached this problem with similar agent based models [10,11]; compared to those, our model places more emphasis on simulating larger populations, computational efficiency, and on code generalisability that allows other researchers to use and develop the code.

We developed the agent-based model (ABM) *OpenABM-Covid19* to simulate an outbreak of COVID-19 in an urban environment. The default population is one million inhabitants with demographic structure based upon UK-wide census data, and household size and age-structure matched to data from the UK 2011 Census survey (for example, older people tend to live together and young children tend to live with younger adults).

On a daily basis all individuals in the model move between networks representing households and either workplaces, schools, or regular social environments for older people. Individuals also interact through random networks representing public transport, transient social gatherings etc. Membership of each type of network is determined by age, giving rise to age-assortative mixing patterns. Network parameters are chosen such that the average number of interactions match age-stratified data reported in[12][12]. The number of daily interactions in random networks is drawn from a negative binomial distribution, allowing for rare super-spreading events.

Infections are seeded in the population and spread through the networks. Biological and epidemiological characteristics of COVID-19 disease have been derived from the scientific literature. The model takes into account asymptomatic infections and different stages of severity, and includes the simulation of hospitalisations and ICU admissions. Since symptoms, disease progression and infectiousness are highly age-dependent, disease pathways in the model are age-stratified.

The ABM was developed to simulate different non-pharmaceutical interventions including lockdown, physical distancing, self-isolation on symptoms, testing and contact tracing. Modelling contact tracing requires the model to keep a record of previous interventions for a set number of days. A variety of contact tracing algorithms are included in the ABM, including tracing on symptoms and/or after a positive test, notifying first-degree contacts only or second-degree contacts as well, testing of traced contacts, and imperfections in test-trace-isolate programmes such as delays, missed contacts and partial compliance. The model reports both aggregated data, such as incidence, tests required, individuals quarantined for various reasons etc., and individual data such as transmission relationships.

OpenABM-Covid19 is available on Github (https://github.com/BDI-pathogens/OpenABM-Covid19), including model documentation, dictionaries for input parameters and output files, over 200 tests in a consistent testing framework used in model validation, and examples for running the model. The core of the model is implemented in the C language for speed; however, the model is run via Python using a SWIG-interface. This interface allows for dynamic intervention strategies to be modelled, as well as providing full transparency about the state of the model. This manuscript was prepared using v0.3 of the model (commit number d14351e) and code for reproducing all figures in this manuscript from model output are publicly available online (https://github.com/BDI-pathogens/OpenABM-Covid19-model-paper).

OpenABM-Covid19 enables simulation of interventions to help policymakers determine the best options to suppress the COVID-19 epidemic in various settings. Default demographic parameters were chosen to reflect the UK and fit well to the UK epidemic after calibration; however, all parameters of the model can be changed by the user.

## 2. Demographics

Within the ABM, individuals are categorised into nine age groups by decade, from “0-9 year ” to “80+ years “. Decades were used because of the strong age-structure of the disease progression. By default, the demographics of the ABM are set to UK national data for 2018 from the Office of National Statistics (ONS). The proportion of individuals in each age group is the same as that specified by the population level statistics in Supplementary Table 1. Since we only consider simulating the epidemics up to a year, we do not consider changes in the population due to births, deaths due to other causes, and migration.

## 3. Interaction network

In each of the interaction networks, individuals are represented as a node. Constant and dynamic connections occur between the nodes in the networks, representing interactions between individuals. The three networks represent different types of daily interactions: household, occupation, and random (Figure 1). The interaction networks have two roles in the ABM. First, the infection can be transmitted between two individuals on a day that they interact. Second, the interactions for each individual are stored and can be used for contact tracing. The membership of different networks leads to age-group assortativity in the interactions. A previous study of social contacts for infectious disease modelling, based on participants being asked to recall their interactions over the past day, has estimated the mean number of interactions that individuals have by age group [12]. We estimate mean interactions by age group by aggregating data (Supplementary Table 2). Figure 2a depicts the resulting distribution of contacts by network and Figure 2b by age.

**Figure 1:**
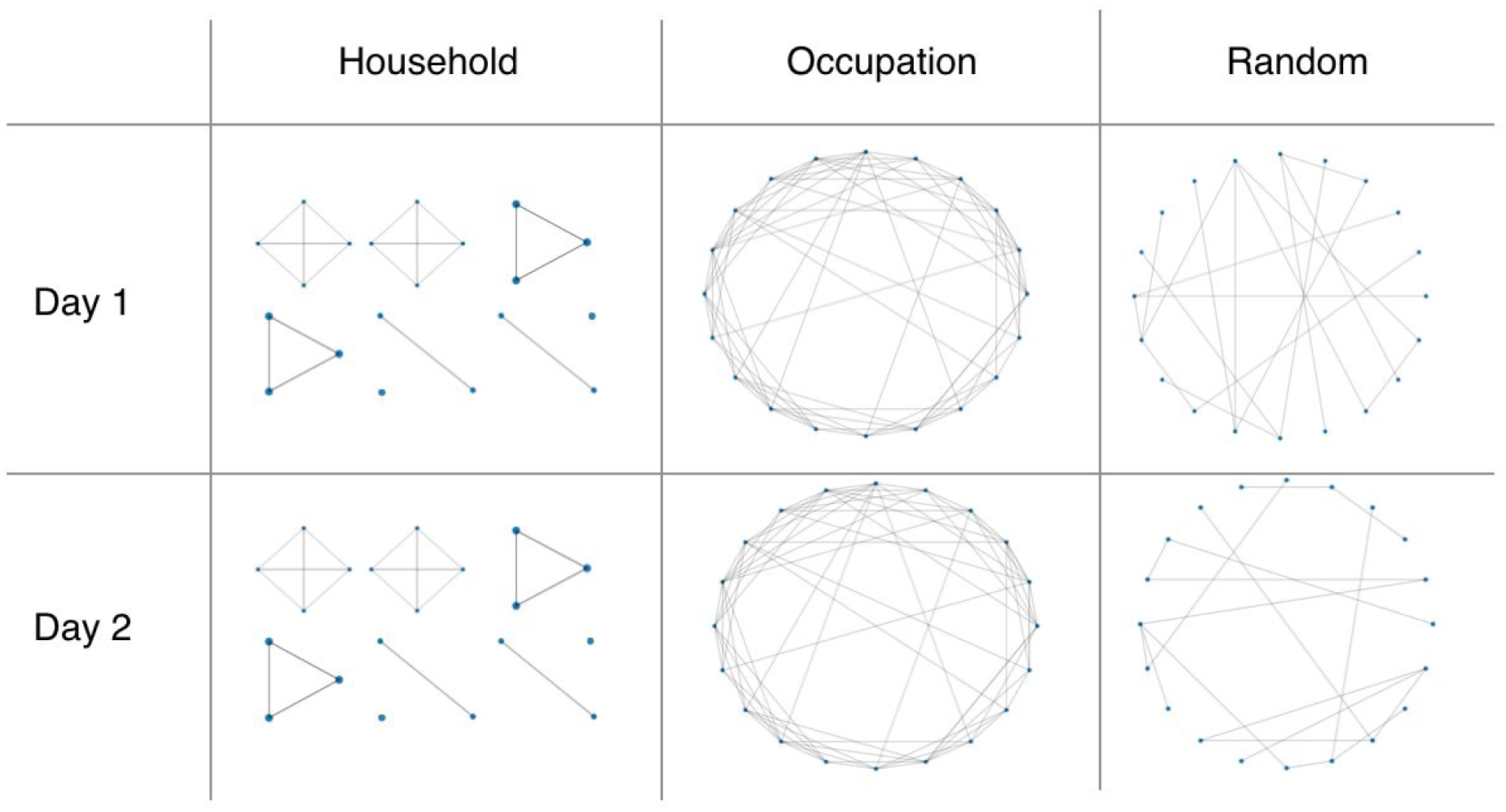
Schematic depiction of the interaction networks. The household and occupation networks are recurrent whereas the random network is transient and rebuilt each day.

**Figure 2:**
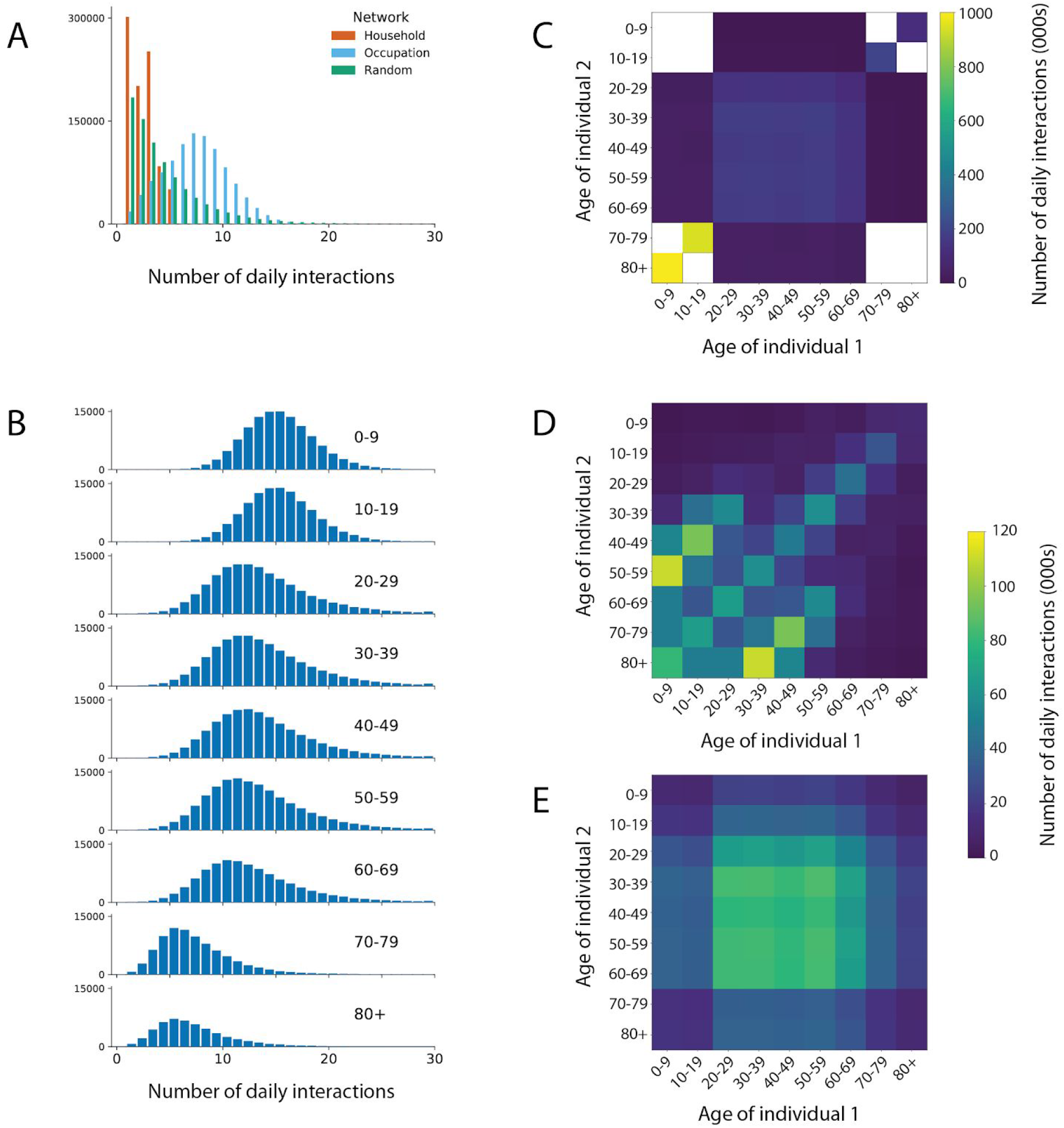
Distribution of daily simulated interactions: A) stratified by network upon which they occur; B) stratified by age group; and stratified by age group of both individuals in the C) occupation, D) household, and E) random networks. Interactions are from the first day of a single simulation in a population of 1 million individuals with UK-like demographics and household structure. Zero counts are shown in white.

Every individual is assigned to live in a single household. The household network is formed by all members of every household interacting with each other every day. The distribution of household sizes is the ONS estimate for the UK in 2018 (Supplementary Table 1). In addition to the household size, the mix of ages in households is important since multi-generational households provide a path by which the infection can be transmitted from young to old. To model this we used a reference panel of 10,000 households taken by down-sampling the UK-wide household composition data from the 2011 Census produced by the ONS. The overall household structure was generated by sampling from the reference household panel with replacement and using rejection-sampling to match the aggregate statistics for the age demographics and household size.

Each individual is also a member of a recurring occupation network to model school, workplace or social networks. The occupation networks are modelled as small-world networks [13]. The network has a fixed set of connections between individuals, and each day a random subset (50%) of these connections are chosen as the interactions between individuals. When constructing the occupation networks, the ABM ensures the absence of overlaps between the household interactions and the local interactions on the small-world network. For children, there are separate occupation networks for the 0-9 year age group (i.e. nursery/primary school) and the 10-19 year age group (i.e secondary school). On each of these networks we introduce a small number of adults (1 adult per 5 children) to represent teaching and other school staff. Similarly for the 70-79 year age group and the 80+ year age group we created separate networks representing daytime social activities among elderly people (again with 1 younger adult per 5 elderly people to represent some mixing between the age groups). All remaining adults (the vast majority) are part of the 20-69 network. Due to the difference in total number of daily interactions, each age group has a different number of interactions in their occupation network. Parameters and values corresponding to the occupation network are shown in Supplementary Table 3.

In addition to the recurring structured networks of households and occupations, we include random interactions. These are drawn randomly each day, independent of previous connections. The number of random connections an individual makes is the same each day (in the absence of interventions), drawn at the start of the simulation from an over-dispersed negative-binomial distribution. This variation in the number of interactions introduces some “super-spreaders ” into the network who have many more interactions than average. The mean numbers of connections were chosen so that the total number of daily interactions matched that from a previous study of social interaction [12]. The number of random interactions was chosen to be lower in children in comparison to other age groups. Interactions in the random network are listed in Supplementary Table 4.

## 4. Infection Dynamics

The infection is spread by interactions between infected (source) and susceptible (recipient) individuals. The rate of transmission is determined by three factors: the infectiousness of the source, the age-dependent susceptibility of the recipient, and the type of interaction, i.e. on which network it occurred.

Infectiousness varies over the natural course of an infection, i.e. as a function of the amount of time the source has been infected, *τ*. Infectiousness starts at zero at the point of infection (*τ* = 0), increases to a peak at an intermediate time, and decreases to zero a long time after infection (large *τ*). Following [7], we took the functional form of infectiousness to be a scaled gamma distribution. We chose the mean and standard deviation as intermediate values between different studies [7,14,15]. Once infected, we split individuals into three groups based upon the eventual severity of the disease: asymptomatic, mild symptomatic and moderate-severe symptomatics. The level of infectiousness depends upon the eventual severity of the disease, i.e. pre-symptomatic individuals who go on to develop moderate-severe symptoms are more infectious than those who go on to develop mild symptoms. By default, the overall infectiousness of asymptomatic individuals and individuals with mild symptoms, is 0.33 and 0.72 times that of individuals with moderate-severe symptoms respectively [16].

An example of how transmissions can be stratified by the infection status of the source and the age of both source and recipient is depicted in Figure 3. In this simulation of an uncontrolled epidemic, most transmissions occur from pre-symptomatic individuals with mild disease who are more numerous than individuals who go on to develop severe disease, followed by symptomatic individuals with mild disease. Interventions that reduce the rate of growth of transmission will change the relative contributions of different symptomatic stages.

**Figure 3:**
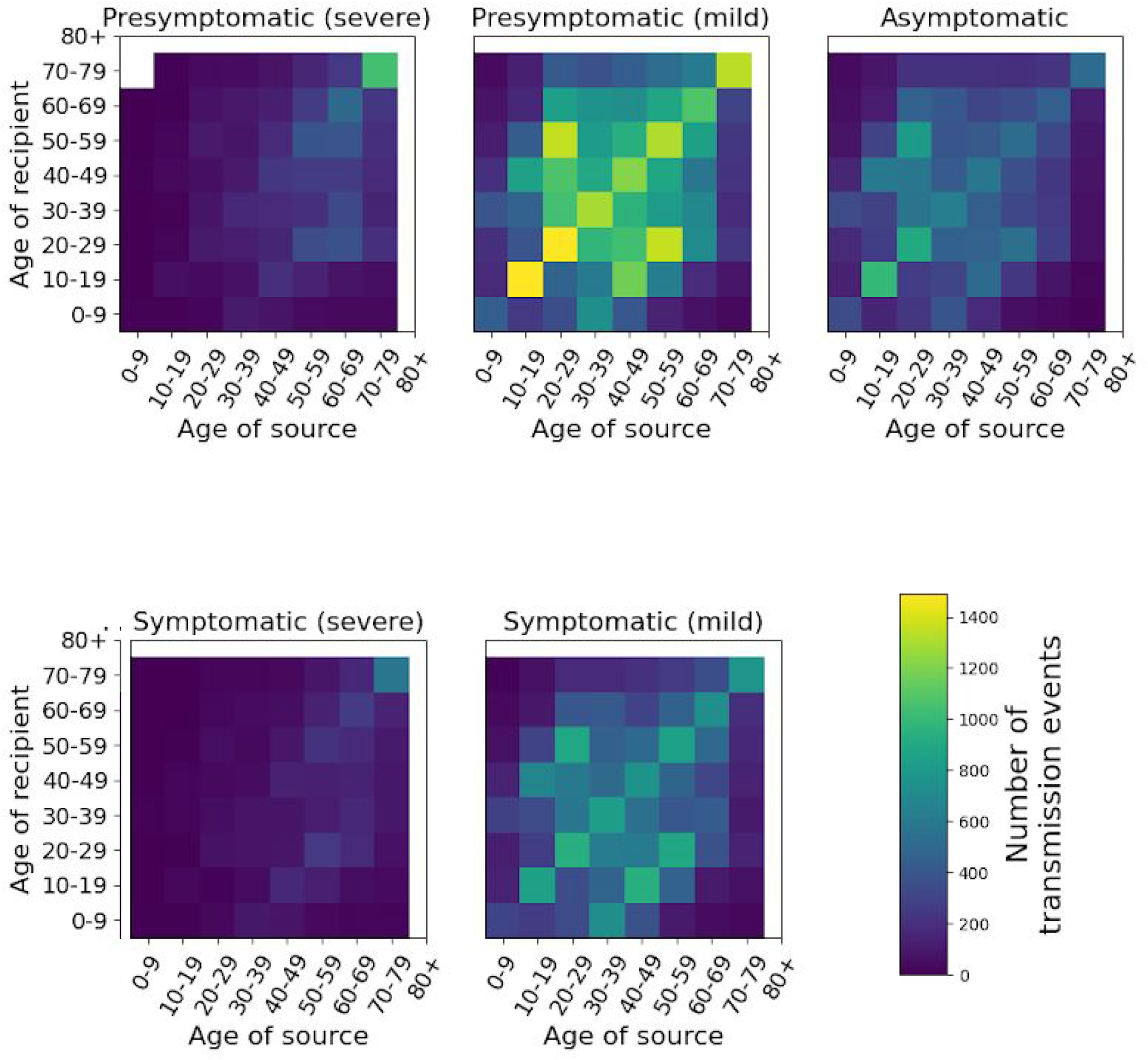
Summary of transmission events from a single simulated uncontrolled epidemic stratified by age of both source and recipient and by infectious status of the source.

The susceptibility of the recipient to infection is modelled with a scale factor dependent on the recipient ‘s age. To calibrate these factors, we identified studies of whether or not transmission occurred from index cases to monitored close contacts [17–24]. Lower probability of infection in children was reported in almost all studies, including that of Zhang et al [17] which observed more infections than the rest of the studies combined, with consistent adjustment for other covariates of transmission risk. We used the susceptibility by age of Zhang et al., interpolated to match our ten-year age categories. The merged data and fit are shown in Supplementary Table 5.

Finally, we model the type of interaction, i.e. on which network the interaction took place. Whilst we do not have data on the length of interactions, interactions which take place within a person ‘s home are likely to be closer than other types of interactions leading to higher rates of transmission. This is modelled using a scale factor, which is 2 by default. Combining all effects, we model the hazard rate per interaction at which the virus is transmitted by

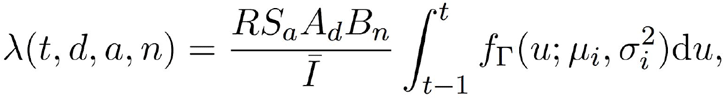

where *t* is the time since the source was infected; *d* indicates the disease severity of the source (asymptomatic, mild, moderate/severe); *a* is the age of the recipient; *n* is the type of network where the interaction occurred; *I* is the mean number of daily interactions; f_Γ_(*u; μ, σ*^*2*^) is the probability density function of a gamma distribution; *μ*_*i*_ and *σ*_*i*_ are the mean and width of the infectiousness curve; *R* scales the overall infection rate; *S*_*a*_ is the relative susceptibility of the recipient based on age; *A*_*d*_ is the relative infectiousness of the source based on disease severity; *B*_*n*_ is the scale factor for the network on which the interaction occurred. Supplementary Table 6 contains the values of the parameters used in simulations. The transmission hazard rate λ is converted to a probability of transmission via *P* = 1 − *e*−λ. The epidemic is seeded by randomly infecting individuals on the day before the simulation starts.

### 5. Natural History of Infection

Upon infection, an individual enters a disease progression cascade where the outcome and rates of progression depend on the age of the infected person. The disease state transitions are shown in Figure 4 and the model parameters in Supplementary Tables 7 and 8.

**Figure 4:**
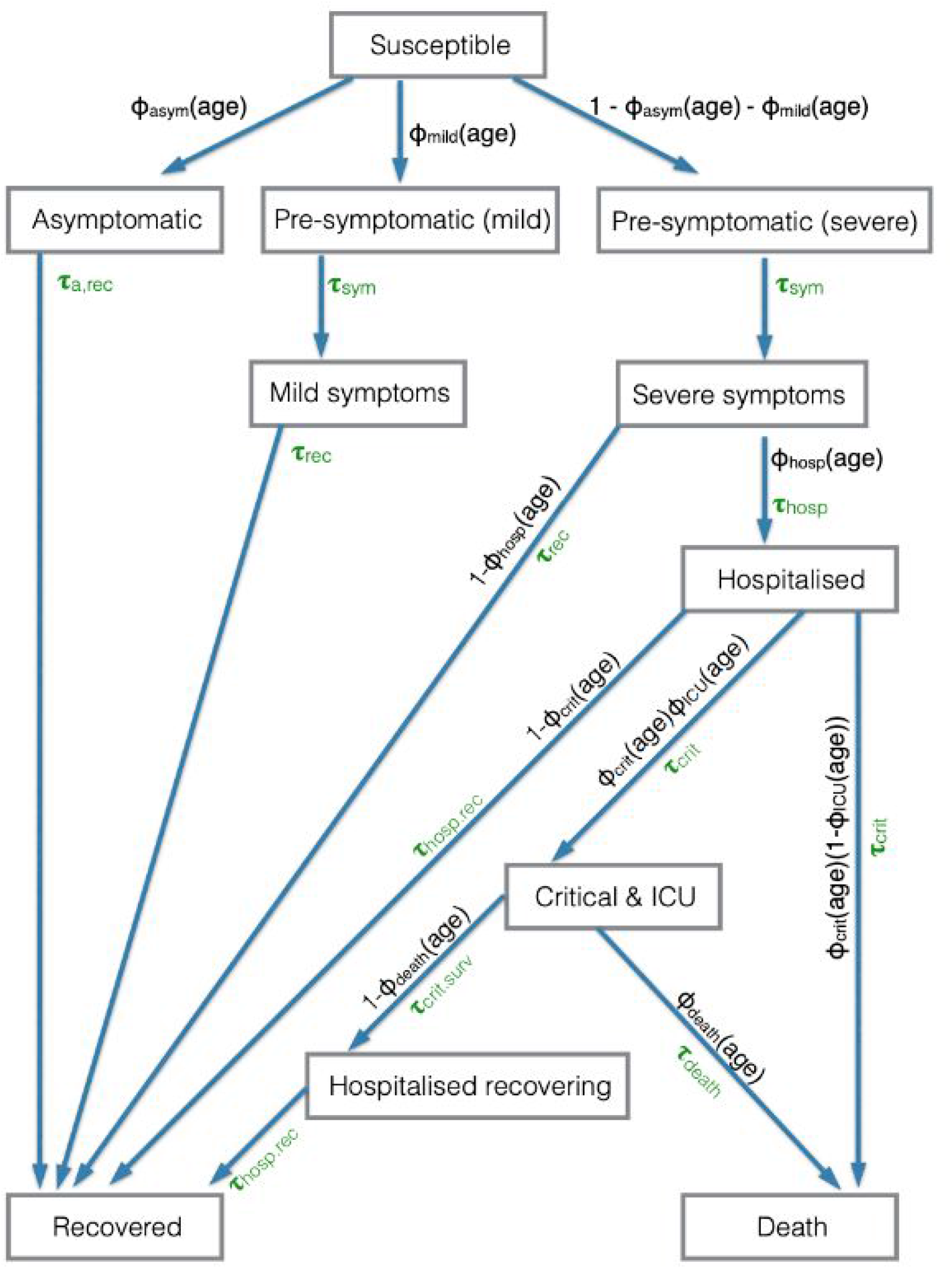
The disease status of an individual, and the probability and time distribution of transitions. The Φ_xxx_(age) variables are the probability of transition to a particular state when there is a choice, where the probability depends upon the age of the individual. The *τ*_xxx_ are the gamma distributed variables of the time taken to make the transition.

A fraction Φ_asym_(age) of individuals are asymptomatic and do not develop symptoms, a fraction Φ_mild_(age) will eventually develop mild symptoms, and the remainder develop moderate/severe symptoms. Each of these proportions depend on the age of the infected individual (Supplementary Table 7). Those who are asymptomatic are infectious at a lower level (see Infection Dynamics section) and will move to a recovered state after a time *τ*_a,rec_ drawn from a gamma distribution.

Once an individual is recovered the model allows immunity to wane through time using two parameters: a fixed period for which every individual must wait, *τ*_waning-shift_, and then a geometric distribution of waiting times until individuals become susceptible, parameterised by its mean *τ*_waning-mean_. By default, the model assumes *τ*_waning-shift_ to be 10,000 days (essentially no waning immunity). During this waiting period, infection is assumed to be completely immunising (recovered individuals cannot be reinfected).

Individuals who will develop symptoms start by being in a pre-symptomatic state, in which they are infectious but have no symptoms. The pre-symptomatic state is important for modelling interventions because individuals in this state do not realise they are infectious and therefore will not self-isolate based on symptoms to prevent infecting others. Individuals who develop mild symptoms do so after time *τ*_sym_ and then recover after time *τ*_rec_ (both drawn from gamma distributions). The remaining individuals develop moderate/severe symptoms after a time *τ*_sym_drawn from the gamma distribution.

Whilst most individuals recover without requiring hospitalisation, a fraction Φ_hosp_(age) of those with moderate/severe symptoms will require hospitalisation. This fraction is age-dependent. Those who do not require hospitalisation recover after a time *τ*_rec_ drawn from a gamma distribution, whilst those who require hospitalisation are admitted to hospital after a time *τ*_hosp_, which is drawn from a shifted Bernoulli distribution. Among all hospitalised individuals, a fraction Φ_crit_(age) develop critical symptoms and require intensive care treatment, with the remainder recovering after a time *τ*_hosp_,_rec_ drawn from a gamma distribution. The time from hospitalisation to developing critical symptoms, *τ*_crit_, is drawn from a shifted Bernoulli distribution. Of those who develop critical symptoms, a fraction Φ_ICU_(age) will receive intensive care treatment. For patients receiving intensive care treatment, a fraction Φ_death_(age) die after a time *τ*_death_ drawn from a gamma distribution, with the remainder leaving intensive care after a time *τ*_crit,surv_. Patients who require critical care and do not receive intensive care treatment are assumed to die upon developing critical symptoms. Patients who survive critical symptoms remain in hospital for *τ*_hosp,rec_ before recovering. The age-dependent infection fatality ratio (IFR) is depicted in Figure 5; other age-dependent outcomes in Supplementary Figure 1. Supplementary Figure 2 shows the corresponding waiting time distributions.

**Figure 5:**
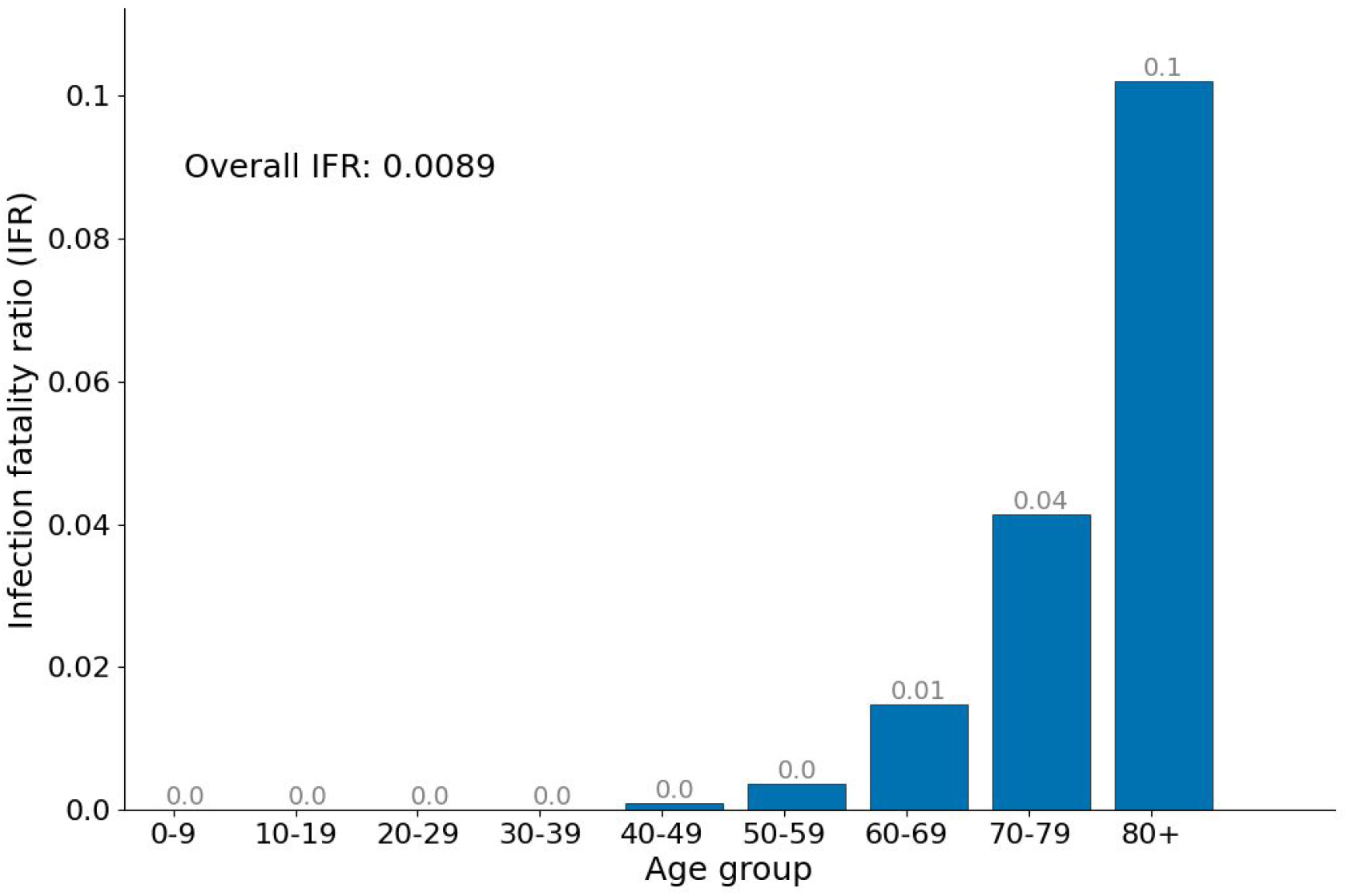
Age-stratified infection fatality ratio (IFR) as output from a single simulation in a population of 1 million with UK-like demography and with a lockdown when prevalence reached 2%. Grey numbers on each bar show the IFR within each age group.

Main outputs of the model include the number of infected individuals, hospitalisations, ICU admissions and deaths (Figure 6). Additional outputs are the number of people in quarantine and the number of tests required, which is of particular interest when comparing different interventions. Transmissions can be depicted according to their type (pre-symptomatic, symptomatic and asymptomatic). The model provides a good fit to UK data (Figure 6).

**Figure 6:**
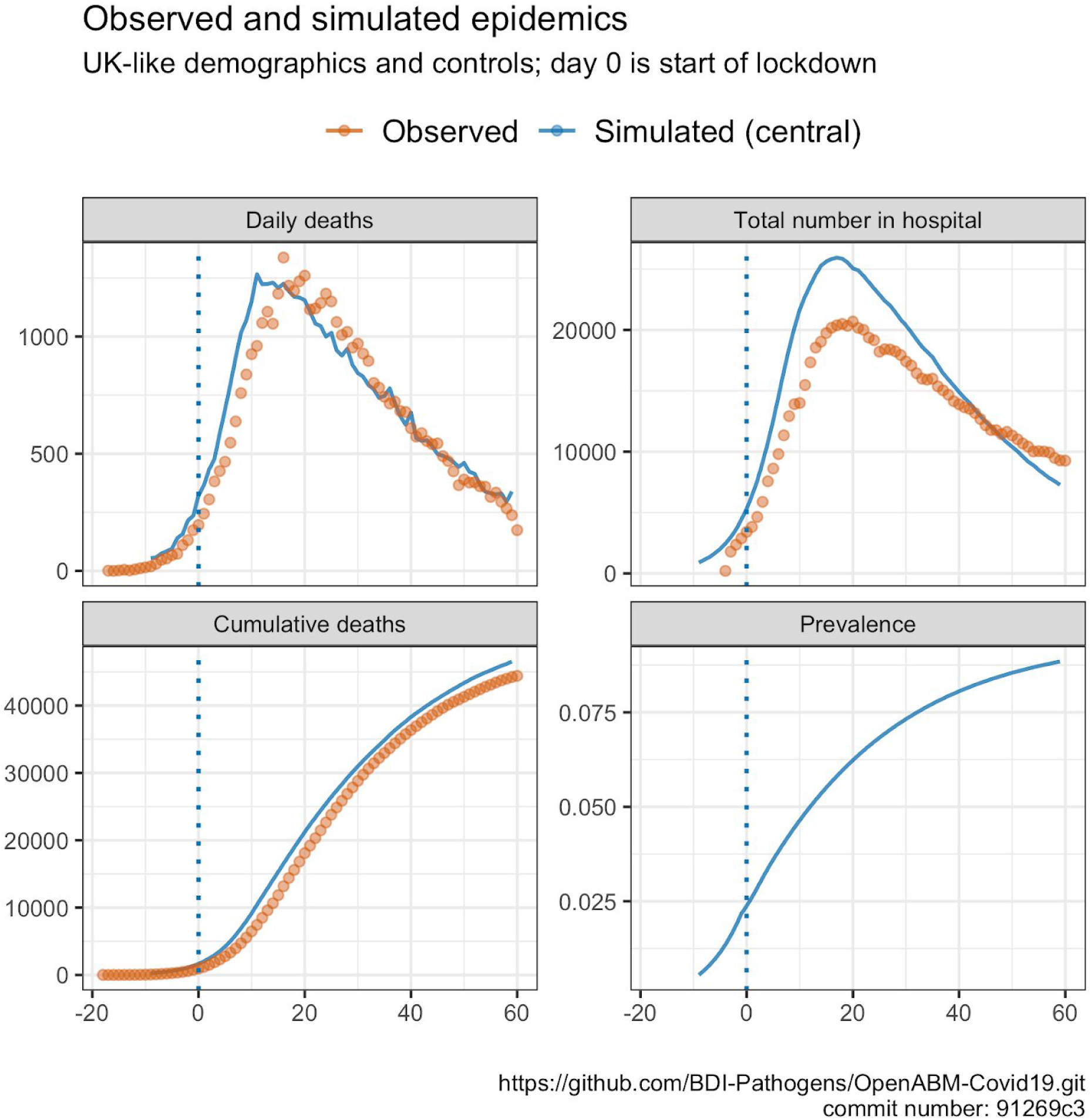
Example of model outputs from a single simulation in a population of 65 million individuals with UK-like demographics and control interventions. Day 0 is the beginning of lockdown (23rd March). Overlaid data are provisional counts of the number of deaths involving the coronavirus (COVID-19) registered in England and Wales (accessed on 5th June 2020), and people in hospital (UK) from UK government press conference 5th June 2020. Simulations are not calibrated to hospitalisation data, only shown for completeness.

## 6. Non-Pharmaceutical Interventions

OpenABM-Covid19 can model a range of non-pharmaceutical interventions (NPIs). Given the many types of intervention and interest in introducing them at different times, the interventions are controlled in the simulation dynamically through the Python interface. This allows for policy interventions to be applied in response to change in the growth of the epidemic (e.g. stricter policies such as lockdown can be applied when prevalence is above a threshold). Below we give brief descriptions of the interventions and sample Python code is given in the Supplementary Materials with links to Jupyter Notebooks. All model parameters involved with NPIs are given in Supplementary Tables 10 and 11.

1. **Self-isolation upon symptoms**: a proportion of individuals self-isolate upon developing symptoms. Self-isolation is modelled by stopping interactions on the individual ‘s occupation network and greatly reducing their number of interactions on the random network. The default time for self-isolation is 7 days with a daily dropout. The ABM contains the option to quarantine everybody within the household of the symptomatic individual. The ABM also considers individuals without COVID-19 who develop flu-like symptoms. Supplementary Figure 4a is a Jupyter Notebook demonstrating how self-isolation upon symptoms reduces the rate of spread of the infection.
2. **Hospitalisation:** once admitted to hospitals, a patient immediately stops interacting with the household, occupation and random networks. We do not model interactions within hospitals, but will add this in future work.
3. **Lockdown**: is modelled by reducing the number of interactions that people have on their occupation and random networks (by default by 71%). Additionally, given that during lockdown people stay at home, the transmission rate for interactions on the household network is increased by a factor of 1.5. Supplementary Figure 4b is a Jupyter Notebook demonstrating the rapid reduction in new infections when a lockdown is imposed. The impact of lockdown on the reproduction number, R, is given in Supplementary Figure 5 and an animation showing the age-stratified detail breakouts is in Supplementary Figure 6.
4. **Shielding**: contact reductions can be applied to certain age groups only. For example, given that fatality ratio is highly skewed towards the over 70s, we have the option of applying a reduction in contacts to this demographic group only. Supplementary Figure 4c is a Jupyter Notebook demonstrating how new infections can be kept low in a shielded group.
5. **Physical distancing**: measures such as physical distancing and mask wearing will reduce the probability of transmission in certain types of interactions (i.e. random interactions but not household interactions). The ABM allows for this to be modelled by allowing for the network-specific transmission multipliers to be adjusted during a simulation. Supplementary Figure 4d is a Jupyter Notebook demonstrating how new infections can be kept low after a lockdown with (extreme) social distancing measures.

### 7. Testing and Contact Tracing

OpenABM-Covid19 is able to model contact tracing (both manual and digital) and how it operates with or without an integrated testing system. The model contains many of the real-world imperfections which affect test and contact tracing programmes, such as test sensitivity and specificity, delays in testing and contact tracing, incomplete coverage, failure to recall contacts, contact tracer resource limitations and impartial adherence to quarantine requests. It also has the ability to model recursive contact tracing with and without testing. Below we give descriptions of the test and contact tracing features, with sample code given in the Supplementary Materials along with links to Jupyter Notebooks.

1. **Testing for SARS-CoV-2 infection**: Testing can occur in both the community and hospital (where an immediate clinical diagnosis is allowed). Tests are assumed to be sensitive from 3 days post-infection to 14 days post-infection with a sensitivity of 80% and specificity of 99%. For community testing, delays can be introduced for ordering a test and for receiving the test result. Testing of an individual in the community is triggered by reporting symptoms and can also be triggered by being contact traced. Supplementary Figure 4e demonstrates the importance in quick testing if self-isolation only occurs after a positive test (as opposed to on symptoms).
2. **Digital Contact Tracing:** Contact tracing is vital to control epidemics with a high level of pre-symptomatic transmission. A variable fraction of individuals in each group can be assigned to have the app. Ownership of smartphones is based on age-stratified OFCOM data (Supplementary Figure 3 and Supplementary Table 9). Digital contact tracing can only occur between two app users. Digital proximity sensing is likely to miss some interactions, so when contact tracing a number of interactions are randomly dropped. For contact tracing, the model takes into account all interactions the individual has had with other app-users for the past seven days which have not been dropped. The model can simulate different app-based contact tracing algorithms. The app can send out notifications with the request to quarantine based on symptoms, or based on a positive test result of the index case. It can ask the household members of the index case and/or household members of the contacts to quarantine and also send out notifications deeper into the network if desired. It can request tests for contacts of index cases if desired. Supplementary Figure 4f demonstrates how digital contact tracing following rapid testing can prevent a second wave even when the average uptake is at only 50% of the total population.
3. **Manual Contact Tracing**: Manual contact tracing works in a similar way to digital contact tracing with a few key differences. First, since it does not rely on an individual being a smartphone user, it can originate from anybody who tests positive (particularly important in the elderly where smartphone usage is lower). However, since the identification of interactions relies on the index case recalling them, only a fraction of actual interactions are traced. In particular, the fraction of interactions recalled depends on the type of interaction (i.e. occupation based interactions are more likely to be recalled than random interactions). Manual contact tracing only occurs after a delay following a positive test, to account for contact tracers contacting both the index and traced individuals. Finally, during a peak in the epidemic the amount of contact tracing required increases and risks overwhelming a manual contact tracing service. Therefore the model contains constraints on the total number of interviews that contact tracers can perform on a single day. Supplementary Figure 4g demonstrates how a well-staffed manual contact tracing following rapid testing can lessen a second wave.
4. **Quarantine**: Contact traced individuals can be asked to quarantine (default 14 days) either because they are directly traced or because they are a household member of somebody who has been traced. Like self-isolation, quarantine is modelled by stopping interactions on the workplace network and greatly reducing the number of interactions on the random network. The model includes a daily dropout rate to simulate imperfect adherence. Quarantine can be ended if the index case later tests negative (after tracing based upon their symptoms), or if the quarantined individual tests negative.

### 8. Implementation details

The core of OpenABM-Covid19 is coded in C using an object-oriented coding style. The code is written in a modular manner to ease readability and encourage extension of the code base. It is open source and is being actively developed by multiple teams. The model uses the GNU Scientific Library (GSL) for mathematical functions, statistical distributions, and random number generation [25] and so any distribution or function available within the GSL can be easily incorporated into the model (for instance in modelling waiting-time distributions). Memory is pre-allocated at the start of the simulation for efficiency.

An important feature of the implementation is the Python interface using SWIG. Running the model via Python allows for complex dynamic interventions strategies to be easily modelled (see examples in Supplementary Figure 4a-h). All states of the model (e.g. transmission events, interactions, individual characteristics) are exposed in Python, which gives full transparency to the results of the model. For example, Supplementary Figure 4h is a Notebook showing how to calculate the relative personal protective effect of for app users versus non-app users when digital contact tracing is used. Python is also a ubiquitous language amongst data scientists, and the interface allows them to fully interact with the model whilst keeping the high speed and memory performance of C.

The model codebase includes over 200 tests used to validate the model. Each test ensures an expected output from the model is realised for a specified set of input parameters. Tests are written in a consistent manner, using the pytest framework. All tests are automatically run when new contributions to the codebase are made. Tests vary input parameters ensuring that expected behaviour of the model is realised across a wide range of input parameter values. Tests cover a range of domains including: disease dynamics, infection and transmission dynamics, non-pharmaceutical interventions, network construction, the C and Python interface, the waiting time distributions, file concordance across the multiple output files from the model, and non-disease related demographics.

Performance. The ABM for 1 million individuals takes approximately 3s per day to run and requires 5Gb of memory (reduced to 1.7Gb if contact-tracing is disabled) on a 2015 MacBook Pro. Both speed and memory are linear in population size (tested from 100k to 1m). The majority of the CPU usage is spent on rebuilding the daily interaction networks and updating the individual ‘s interaction diaries.

## 9. Discussion

We present OpenABM-Covid19, a COVID-19-specific agent-based model suitable for simulating the epidemic in different settings and assessing non-pharmaceutical interventions, including contact tracing using a mobile phone app. The model is well documented with a simple interface, allowing non-experts to easily evaluate complex dynamic intervention strategies in a few lines of Python code. OpenABM-Covid19 is an open-source project and is easily extensible, with new features already being added by multiple external teams. The model is fully documented and is thoroughly tested in a formal testing framework.

The model was designed to be as parsimonious as possible, with complexity only added when it was essential to model important features of COVID-19 or details of non-pharmaceutical interventions, and with parameters being inferred from published studies. Due to the substantial pre-symptomatic and asymptomatic transmission of the virus, it is necessary to model each individual ‘s normal daily interactions. Further, on developing symptoms or during interventions such as contact tracing, the interaction pattern of individuals change to only include those in the household. We therefore took the decision to model interactions using three social networks (household/occupational/random) with non-pharmaceutical interventions affecting each network differently. Recurring small-world networks were used to model interactions at home and at work, whereas a transient random network was used to model other daily interactions such as on public transport or in shops. The strong association of COVID-19 disease progression with age along with the age assortativity of social networks, led us to using a decade age-structure. The model simulated an urban population of 1 million rather than the population of a whole country to allow realistic estimates for hospitalisation and ICU admission forecasts on a regional level. Large national epidemics will also exhibit meta-population dynamics rather than the spatially unstructured mixing modelled here.

One of the key aims of OpenABM-Covid19 was to model non-pharmaceutical interventions and, in particular, different forms of contact tracing. The model of digital contact-tracing allows for questions such as the role of: testing delays, different quarantine requests, compliance rates, recursive testing, and app uptake to be investigated. The model of manual contact-tracing allows for questions such as resource limitations, partial contact recall and interview delays to be investigated. Importantly, due to the simple Python interface, it is possible for non-experts to simulate all these features and to investigate the effect of applying multiple intervention policies at different stages of the epidemic.

The current version of the model does not currently include events in hospitals, care-home settings, non-hospital deaths, gender/sex of individuals, comorbidities, or any geographical structure apart from that implicit within the three modelled networks. All of these limitations are being currently addressed by collaborators and will become available on the Github repository in the near future.

OpenABM-Covid19 is a versatile tool to model the COVID-19 epidemic in different settings and simulate different non-pharmaceutical interventions including contact tracing. OpenABM-Covid19 is a modular tool that will help scientists and policymakers weigh decisions during this epidemic. Our vision is that, with the help of the world-wide modelling community, it will develop into a family of models for infectious diseases that are at risk of causing pandemics in the future, adding to the international toolkit for epidemic preparedness.

## Data Availability

Data sharing not applicable, no new data generated.

## Notes

### Competing Interest Statement

M.A. and N.W. are employees of Alphabet, Inc., a provider of the Exposure Notification System; no other relationships or activities that could appear to have influenced the submitted work.

### Funding Statement

The study was funded by an award from the Li Ka Shing Foundation to CF. The funders of the study had no role in study design, data collection, data analysis, data interpretation, or writing of the report.

### Author Declarations

This work does not require approval from a IRB.

## References

1. World Health Organization. Coronavirus disease weekly epidemiology report, 6th September 2020. Available: https://www.who.int/docs/default-source/coronaviruse/situation-reports/20200907-weekly-epi-update-4.pdf?sfvrsn=f5f607ee_2

2. Brodeur A, Gray DM, Islam A, Bhuiyan S. A Literature Review of the Economics of Covid-19. 2020. Available: https://papers.ssrn.com/abstract=3636640

3. Metcalf CJE, Morris DH, Park SW. Mathematical models to guide pandemic response. Science. 2020;369:368–369.

4. Anderson RM, Fraser C, Ghani AC, Donnelly CA, Riley S, Ferguson NM, et al. Epidemiology, transmission dynamics and control of SARS: the 2002--2003 epidemic. Philos Trans R Soc Lond B Biol Sci. 2004;359:1091–1105.

5. Adam D. Special report: The simulations driving the world ‘s response to COVID-19. Nature. 2020;580:316–318.

6. Chang SL, Harding N, Zachreson C, Cliff OM, Prokopenko M. Modelling transmission and control of the COVID-19 pandemic in Australia. arXiv preprint. 2020;arXiv:2003.10218. Available: http://arxiv.org/abs/2003.10218

7. Ferretti L, Wymant C, Kendall M, Zhao L, Nurtay A, Abeler-Dörner L, et al. Quantifying SARS-CoV-2 transmission suggests epidemic control with digital contact tracing. Science. 2020;368. doi:10.1126/science.abb6936

8. Kucharski AJ, Klepac P, Conlan AJK, Kissler SM, Tang ML, Fry H, et al. Effectiveness of isolation, testing, contact tracing, and physical distancing on reducing transmission of SARS-CoV-2 in different settings: a mathematical modelling study. Lancet Infect Dis. 2020:30457–6. doi:10.1016/S1473-3099(20)30457-6

9. Hinch R, Probert W, Nurtay A, Kendall M, Wymant C, Hall M, et al. Effective configurations of a digital contact tracing app: A report to NHSX. en In:(Apr 2020) Available here url: https://github.com/BDI-pathogens/covid-19_instant_tracing. 2020.

10. Kerr CC, Stuart RM, Mistry D, Abeysuriya RG, Hart G, Rosenfeld K, et al. Covasim: an agent-based model of COVID-19 dynamics and interventions. medRxiv. 2020. Available: https://www.medrxiv.org/content/10.1101/2020.05.10.20097469v1.abstract

11. Bicher MR, Rippinger C, Urach C, Brunmeir D, Siebert U, Popper N. Agent-Based Simulation for Evaluation of Contact-Tracing Policies Against the Spread of SARS-CoV-2.medRxiv.2020.Available: https://www.medrxiv.org/content/10.1101/2020.05.12.20098970v2.abstract

12. Mossong J, Hens N, Jit M, Beutels P, Auranen K, Mikolajczyk R, et al. Social contacts and mixing patterns relevant to the spread of infectious diseases. PLoS Med. 2008;5(3):e74.

13. Watts DJ, Strogatz SH. Collective dynamics of “small-world ” networks. Nature. 1998;393:440–442.

14. Ma S, Zhang J, Zeng M, Yun Q, Guo W, Zheng Y, et al. Epidemiological parameters of coronavirus disease 2019: a pooled analysis of publicly reported individual data of 1155 cases from seven countries. medRxiv. 2020. doi:10.1101/2020.03.21.20040329

15. Ganyani T, Kremer C, Chen D, Torneri A, Faes C, Wallinga J, et al. Estimating the generation interval for coronavirus disease (COVID-19) based on symptom onset data, March 2020. Euro Surveill. 2020;25. doi:10.2807/1560-7917.ES.2020.25.17.2000257

16. Sun et al. (2020) Personal communication.

17. Zhang J, Litvinova M, Liang Y, Wang Y, Wang W, Zhao S, et al. Changes in contact patterns shape the dynamics of the COVID-19 outbreak in China. Science. 2020;368:1481–1486.

18. Li W, Zhang B, Lu J, Liu S, Chang Z, Peng C, et al. Characteristics of Household Transmission of COVID-19. Clin Infect Dis. 2020. doi:10.1093/cid/ciaa450

19. Luo L, Liu D, Liao X-L, Wu X-B, Jing Q-L, Zheng J-Z, et al. Modes of contact and risk of transmission in COVID-19 among close contacts. medRxiv. 2020. doi:10.1101/2020.03.24.20042606

20. Jing Q-L, Liu M-J, Zhang Z-B, Fang L-Q, Yuan J, Zhang A-R, et al. Household secondary attack rate of COVID-19 and associated determinants in Guangzhou, China: a retrospective cohort study. Lancet Infect Dis. 2020. doi:10.1016/S1473-3099(20)30471-0

21. Wei L, Lv Q, Wen Y, Feng S, Gao W, Chen Z, et al. Household transmission of COVID-19, Shenzhen, January-February 2020. medRxiv. 2020; 2020.05.11.20092692.

22. Chaw L, Koh WC, Jamaludin SA, Naing L, Alikhan MF, Wong J. SARS-CoV-2 transmission in different settings: Analysis of cases and close contacts from the Tablighi cluster in Brunei Darussalam. medRxiv. 2020; 2020.05.04.20090043.

23. Bi Q, Wu Y, Mei S, Ye C, Zou X, Zhang Z, et al. Epidemiology and transmission of COVID-19 in 391 cases and 1286 of their close contacts in Shenzhen, China: a retrospective cohort study. Lancet Infect Dis. 2020;20:911–919.

24. Xu Y, Li X, Zhu B, Liang H, Fang C, Gong Y, et al. Characteristics of pediatric SARS-CoV-2 infection and potential evidence for persistent fecal viral shedding. Nat Med. 2020;26:502–505.

25. Galassi M, Davies J, Theiler J, Gough B, Jungman G, Alken P, et al. GNU Scientific Library Reference Manual (Network Theory Ltd.), 3rd (v1. 12) edition. 2009.

